# Outcomes of Anti-Spike Monoclonal Antibody Therapy in Pregnant Women with Mild to Moderate COVID-19

**DOI:** 10.1101/2021.11.27.21266942

**Authors:** Bright P. Thilagar, Aditya K. Ghosh, Jerome Nguyen, Regan N. Theiler, Myra J. Wick, Ryan T. Hurt, Raymund R. Razonable, Ravindra Ganesh

## Abstract

**Objective:** To evaluate the efficacy and safety of anti-spike monoclonal antibodies (MAb) in the treatment of mild to moderate COVID-19 in high-risk patients who are pregnant.

**Methods:** The database of patients treated with monoclonal antibodies in the Mayo Clinic Midwest region was reviewed for patients who were pregnant at the time of infusion. Manual chart review was performed to collect demographic details as well as COVID course for both the mother and the infant if delivered. The data are presented using descriptive methods.

**Results:** We identified fifty-one pregnant patients with mild to moderate COVID-19 who were treated with MAb (4 with bamlanivimab monotherapy, 3 with bamlanivimab-etesevimab combination, and 44 with the casirivimab-imdevimab combination). No adverse effects were reported, and no patient required COVID-19 related hospitalization. Twenty-nine patients delivered healthy babies, there was one case of intrauterine fetal demise secondary to a congenital Ebstein anomaly (not related to MAb treatment), and twenty-one were uncomplicated pregnancies.

**Conclusion:** MAb infusions were well tolerated in pregnant patients considered at high risk for COVID-19 complications, with no observed adverse effects to mother or fetus. Although preliminary data suggest MAb therapy in pregnancy is safe, further research is recommended to fully assess safety and efficacy in pregnancy.

**TEACHING POINTS:**

- Anti-spike monoclonal antibody therapy is well tolerated in high-risk pregnant patients with mild to moderate COVID-19
- No adverse effects of anti-spike monoclonal antibody administration were observed in either the mother or fetus.

## Introduction

Coronavirus disease 2019 (COVID-19) caused by Severe Acute Respiratory Syndrome Coronavirus-2 (SARS-CoV-2) has been associated with hospitalization and death, particularly in certain high risk patient groups.^1,2^ COVID-19 has been associated with a worse clinical course in pregnant women when compared with non-pregnant women.^3-6^ While the majority of pregnant women with COVID-19 recover without developing severe disease requiring hospitalization, several reports indicate that some may be at increased risk of complications, especially those with concomitant risk factors including age over 35 years, obesity, diabetes mellitus (DM) and hypertension.^6,7^

The anti-spike monoclonal IgG antibody preparations (MAb’s), bamlanivimab with or without etesevimab, the combination of casirivimab and imdevimab, and sotrovimab, have received separate emergency use authorizations (EUA) from the US Food and Drug Administration (FDA) for the outpatient treatment of mild-moderate COVID-19 in adult patients at increased risk of severe outcomes. High risk in this context includes body mass index (BMI) ≥ 25 kg/m^2^, age ≥ 65 years, DM, immunosuppressive disease or therapy, chronic kidney disease, pregnancy, neurologic disease, dependence on medical device, cardiovascular disease, chronic pulmonary disease, or hypertension. These US FDA EUAs were based on separate randomized, placebo controlled clinical trials of bamlanivimab monotherapy, combination of bamlanivimab and etesevimab, combination of casirivimab and imdevimab, and sotrovimab. These randomized controlled trials demonstrated reduction in viral load along with decrease in medically attended visits and hospitalizations among patients who received anti-spike monoclonal antibodies compared to the placebo group.^8-11^

Pregnancy worsens the clinical course of COVID-19 and was listed in the FDA EUA criteria as a condition that renders a patient at high-risk of COVID-19 complications.^12^ However, data regarding use of MAb therapy in pregnant women with COVID 19 is limited as they were not included in the clinical trials. Small case series have reported the safety of MAb in pregnant women.^13,14^ Accordingly, there is hesitancy on the use of the MABs in pregnant women, despite their inclusion in the FDA EUA guidance. Societal guidelines have initially suggested that pregnancy is not a contraindication to potentially effective treatment regimens for COVID-19.^15^ The American College of Obstetrics and Gynecology and the Society of Maternal and Fetal Medicine also state that there is no evidence for or against the use of these products in pregnancy.^16^ These societies further suggested that adult pregnant patients at high risk of progressing to severe COVID-19 and/or hospitalization may be offered MAb therapies after discussion of their potential risks and benefits in a shared decision-making process.^17^ On May 14, 2021, the FDA expanded the EUA to include pregnancy as a qualifying condition for MAb.

In this report, we present a large case series that describes the clinical outcomes of 51 pregnant women with mild to moderate COVID-19, who have consented for the infusion of anti-spike MAb therapy after a shared decision-making process.

## Methods

The Mayo Clinic Monoclonal Antibody Treatment (MATRx) Program started on November 9, 2020 to facilitate the administration of anti-spike MAb therapies based on the FDA EUA directive. The details of this program have been published.^18^ All patients with positive SARS-CoV-2 PCR or antigen test were automatically evaluated for eligibility by the MATRx providers, and all eligible patients were proactively contacted for patient education and consenting for treatment.

Between November 19, 2020 to September 23, 2021, over 10,800 high-risk patients with mild to moderate COVID 19 received anti-spike MAb therapy at outpatient COVID-19 Infusion Therapy Centers of the Mayo Clinic in the Midwest. The choice of MAb therapy was dependent on the available supply at the time of infusion. Bamlanivimab (700-mg) monotherapy, bamlanivimab (700-mg) with etesevimab (1400-mg), casirivimab (1200-mg) with imdevimab (1200-mg), and sotrovimab (500-mg) were available options. The use of bamlanivimab monotherapy was revoked by the US FDA on April 16, 2021 due to variant resistance. All were administered as a single infusion over 30-60 minutes at the dedicated COVID-19 Infusion Therapy Center (ITC). All patients were monitored for adverse reaction during the infusion and for one hour after the infusion. All patients were followed with the Remote Patient Monitoring Program to assess stability of their medical condition, clinical symptoms and adverse effects.

After approval by the Mayo Clinic Institutional Review Board (IRB), the MATRx Registry was queried for patients who received MAb and had a diagnosis of pregnancy.^18^ A total of 86 pregnant patients were identified and their medical records were reviewed and abstracted to collect details such as age, gestational history, gestation at COVID-19 diagnosis, dates of symptom onset and laboratory diagnosis, COVID-19 symptoms, date and type of MAb administration, additional medications, complications, and hospitalization. The data are presented using descriptive methods.

## Results

### Patient characteristics and clinical disease

After excluding patients who had false-positive pregnancy tests and those who received the monoclonal antibodies after delivery, there were fifty-one pregnant women who received anti-spike MAb for mild to moderate COVID-19 during pregnancy (Figure 1). The demographic characteristics of the patient cohort are listed in Table 1. The mean age was 31.1 years old with a range from 22-40 years old. Of these patients, 4 received bamlanivimab monotherapy (prior to April 16, 2021), 3 received the bamlanivimab-etesevimab combination, and 44 received the casirivimab-imdevimab combination. No patient received sotrovimab. MAb was administered to 4 patients during the first trimester, 17 in the second trimester, and 30 in the third trimester, with an overall mean gestational age of 180.1 days. Major co-existing comorbidities were depression (n=21, 41%), migraine (n-17, 33%), obesity with BMI >30 (n=11, 22%), asthma (n=8, 16%), anxiety (n=5, 10%), and DM (n=3, 6%).

**Table 1:** Demographic Characteristics of patients at time of MAb infusion.

| Criterion | Mean (SD) or n(%) |
| --- | --- |
| Age | 31.1 (4.7) |
| Gravida | 3.4 (2.0) |
| Parity | 1.5 (1.3) |
| **Race** |  |
| White | 46 (90%) |
| African American | 1 (2%) |
| American Indian | 1 (2%) |
| Pacific Islander | 1 (2%) |
| Other | 2 (4%) |
| **Ethnicity** |  |
| Not Hispanic/Latino | 47 (92%) |
| Hispanic/Latino | 4 (8%) |
| **Language** |  |
| Arabic | 1 (2%) |
| English | 49 (96%) |
| Spanish | 1 (2%) |
| **Co-Morbidities** |  |
| Depression | 21 (41%) |
| Migraine | 17 (33%) |
| BMI>30 | 11 (22%) |
| Asthma | 8 (16%) |
| Anxiety | 5 (10%) |
| **Infusion** |  |
| Time to Infusion (days) | 2.9 (1.6) |
| Gestational age at infusion (days) | 180.1 (70.2) |

**Table 2:**
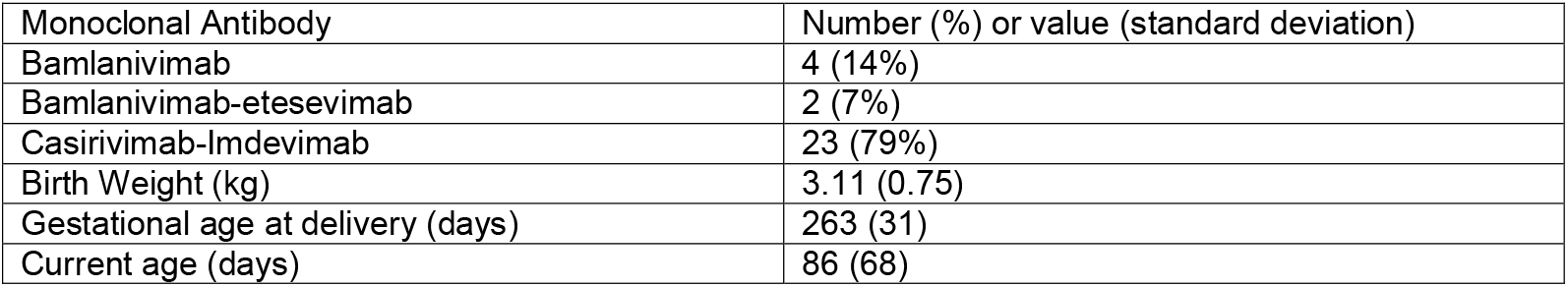
Characteristics of infants delivered after MAb infusion during pregnancy.

**Figure 1:**
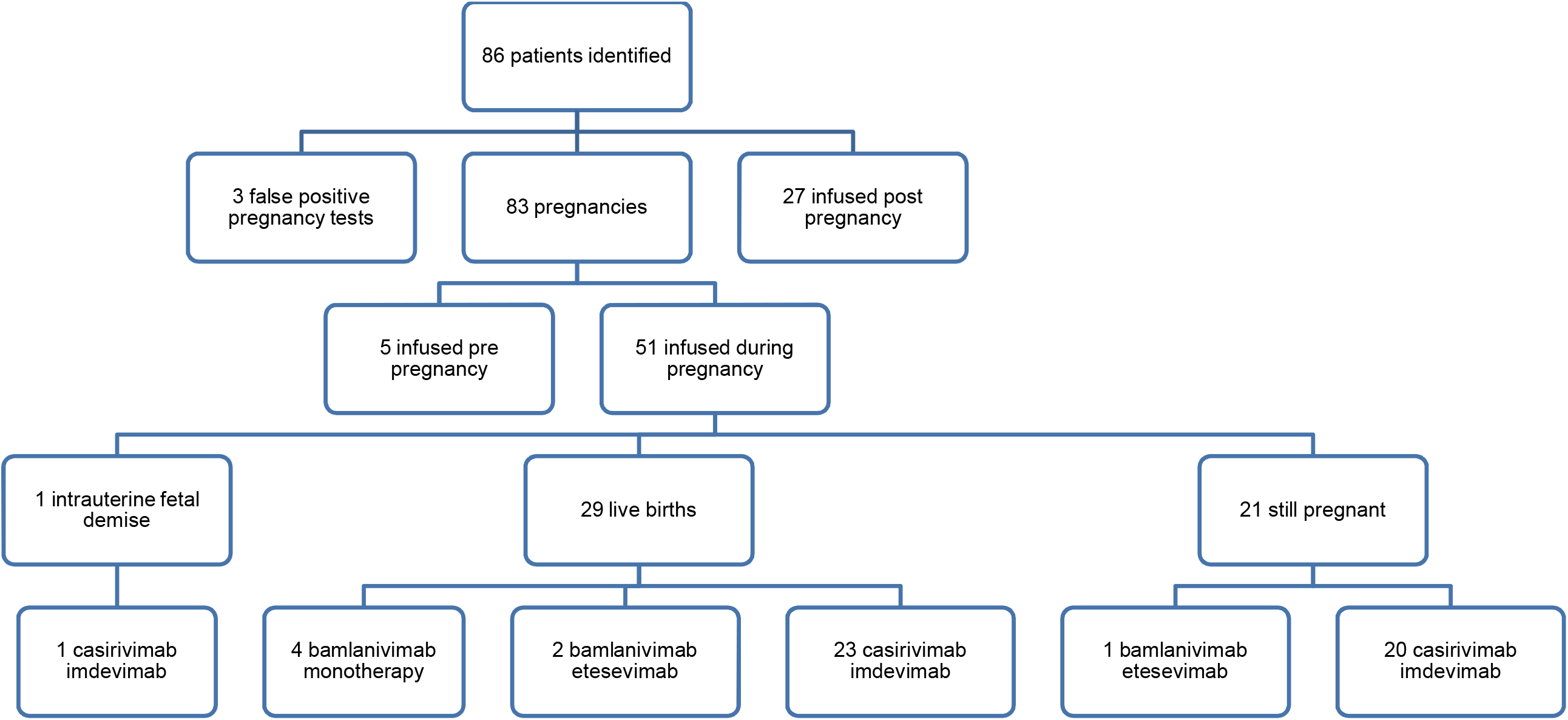
Patient inclusion and characteristics.

In accordance with the FDA EUA guidance, all patients were classified as having mild to moderate COVID-19. Patients with severe illness at the time of clinical presentation were excluded from Mab therapy. The duration from positive COVID-19 PCR test result to the administration of MAb therapy ranged from 1-8 days, with a mean of 2.9 days.

### Treatment effects and outcome

All patients were monitored during and for at least one hour after the Mab infusion. None of the 51 patients experienced immediate adverse effects during or after administration of MAb. None of the patients required any additional COVID-19 directed therapy outside of MAb, such as remdesivir.

After Mab infusion, 10 patients sought care in the Emergency Department (ED) for a variety of reasons with only one related to COVID-19 (dyspnea). The other causes of ED visits not leading to admission were temporally separated from Mab infusion by at least 30 days and included nausea and vomiting of pregnancy, asthma, cough, and post-operative pain.

Four patients were subsequently admitted to the hospital, but these were not likely related to MAb infusion: one was admitted for unrelenting headache needing sphenopalatine ganglion blockade at 42 days after MAb infusion, one was admitted for observation of a transient ischemic attack 71 days after MAb infusion, one was admitted for a flare of Crohn’s disease at 175 days after MAb infusion, and one was admitted for intrauterine fetal demise secondary to Ebstein anomaly. The intrauterine fetal demise secondary to Ebstein anomaly was diagnosed two weeks after receiving Mab at 14 weeks and 5 days of gestational age. This progressed to fetal hydrops and fetal demise which led to dilation and evacuation. The diagnosis and associated complications were deemed to be not associated with MAb therapy after review by the MATRx team and our Obstetrics specialists.

No patient had pregnancy complications at the time of follow up. No case of fetal distress was reported secondary to anti-spike MAb infusion. At the time of this report, 29 patients who had been treated with MAb in pregnancy had delivered, and 21 were still pregnant. The 29 infants delivered were all doing well ranging between 5 and 310 days old (mean age 86 days, mean birthweight 3.11 kg).

## Discussion

We describe the clinical outcomes of fifty-one pregnant patients who received anti-spike MAb for mild to moderate COVID-19. To our knowledge, this is the largest series to date in this population. MAb therapy was generally tolerated well, with no immediate adverse effects or complications related to the infusion. All fifty-one patients had resolution of their COVID-19 symptoms without progression to severe disease requiring hospitalization, supplemental oxygen therapy or additional antiviral therapies. Only one patient had an ED visit that could be related to COVID-19. At the time of this report, twenty-nine patients have delivered healthy infants without complication while twenty-one are still pregnant without reported complications. There was one case of intrauterine fetal demise secondary to congenital Ebstein anomaly which led to hydrops fetalis and intrauterine death. This was likely not related to the anti-spike monoclonal antibody infusion; two cases of hydrops fetalis leading to intrauterine death have also been reported as a complication of COVID-19 likely due to vertical transmission of SARS-CoV-2.^19,20^

One of the concerns for experimental therapies in pregnant women is the potential for the drugs to cross into the fetal environment. The anti-spike IgG-subtype MAb may cross the placenta, although the level reaching the fetus is unknown. Since the anti-spike MAb is directed against the virus, and not directed towards human antigens, it is theoretically safe for administration in pregnancy. However, the clinical data to support this theoretical assumption is not currently available for COVID-19 MAbs, as pregnant patients were not included in the investigational clinical trials. Review of the literature suggests that other forms of MAb have been given and are reportedly safe in pregnant women. Tocilizumab, a humanized MAb that targets interleukin-6, has been used to counteract the inflammatory state of COVID-19 in some pregnant women without reported detrimental effects.^21,22^ In another report of 23 patients from the German Multiple Sclerosis and pregnancy registry who received other MAb (natalizumab and ocrelizumab) during lactation and the third trimester of pregnancy, there was no impact on infant health.^23^ Despite these anecdotal experiences of potential safety profile, it is important to document clinical experiences with these products, in order to better guide their use. It is in this context that we are reporting our experience in treating fifty-one pregnant women. Our report suggests that COVID-19 MAbs appear to be safe and well tolerated during pregnancy, with no untoward effects to the mother and the fetus. Nonetheless, we emphasize the need to discuss these therapeutic options with every pregnant patient in a shared decision-making process.

The risk of adverse pregnancy outcomes is increased in pregnant women with severe and critical COVID-19. Accordingly, the US FDA had suggested providing MAbs to pregnant patients with COVID-19 in order to prevent progression to severe and critical disease. The current study reports that the outcomes of anti-spike MAb infusion in fifty-one pregnant women was favorable, since none of the patients progressed to severe or critical illness. It is possible that MAb mitigated the progression to severe disease and thus, this may have led to the favorable outcomes among our cohort. However, we emphasize that this data should only serve as preliminary safety data in this population, and further studies will be needed to confirm our observations. Of the three MAb therapies infused in this group, two (bamlanivimab-etesevimab and casirivimab-imdevimab) remain effective against circulating variants. Specifically, in our sample, 38 pregnant women were infused after July 1, 2021 when the delta variant gained predominance in the Midwest.^24^ Over this time period, our center was preferentially using casirivimab-imdevimab with significant reduction in hospitalization.^24^ Notably, this study did not include patients treated with sotrovimab, as this product was not available at our infusion facilities when any of the fifty-one pregnant patients presented for their MAb therapies.

In conclusion, this case series is amongst the earliest reports of the use of anti-spike MAb during pregnancy. Importantly, all patients had an uncomplicated course of MAb infusion. No patient progressed to severe COVID-19 or required COVID-19 related hospitalization or had demonstrable adverse fetal effects from the therapy. The single case of intrauterina fetal demise due to Ebstein anomaly was deemed not related to MAb therapy. Despite these reassuring findings from a limited number of patients, further research with larger population of diverse patients and the use of combination therapy is warranted. In addition, long term follow-up on the patients and of their newborns will be important. To date, in our series, twenty-nine infants have been delivered and are doing well with age up to 310 days.

## Data Availability

All data produced in the present study are available upon reasonable request to the authors

## Notes

### Competing Interest Statement

AKG, JN, RNT, MJW, BPT, and RG have no financial disclosures.
RRR is principal investigator of clinical trials on COVID-19 treatment funded by Gilead (remdesivir), Regeneron (sarilumab, casirivimab-imdevimab), Roche (tocilizumab), and research on Monoclonal Antibody Therapy funded by the Mayo Clinic. RTH is a consultant for Nestle Nutrition

### Funding Statement

This study was funded by the Mayo Clinic

### Author Declarations

Mayo Clinic IRB gave ethical approval for this work

